# Uptake and feasibility of task-shifting of Xpert MTB/RIF Ultra testing from laboratory technicians to nurses to increase access and reduce time to results: a multi-country mixed method research

**DOI:** 10.1101/2025.03.24.25324527

**Authors:** Eden Masama Ngu, Basant Joshi, Bandana Bhatta, Minh Huyen Ton Nu Nguyet, Manon Lounnas, Rodney Kaitano, Marcelle N’Guessan, Kunda Kasakwa, Emelva Manhiça, Song Yin, Jacob Mugisha, Timothée Ouassa, Jean-Voisin Taguebue, Raoul Moh, Guillaume Breton, Juliet Mwanga-Amumpere, Laurence Borand, Chishala Chabala, Celso Khosa, Eric Wobudeya, Olivier Marcy, Joanna Orne-Gliemann, Maryline Bonnet, the TB Speed study group

## Abstract

**Introduction:** Task-shifting of tuberculosis (TB) rapid molecular testing from laboratory technicians to nurses could help decentralizing TB diagnosis at Primary Health Centers (PHC) and allow in-ward testing to shorten treatment decision for very sick patients. We assessed the feasibility of XpertMTB/RIF Ultra (Ultra) testing on nasopharyngeal aspirate (NPA) by nurses in children with presumptive TB at PHCs and in hospitalized children with severe pneumonia within the TB-Speed project in seven countries.

**Methods:** Of 23 PHCs and 15 paediatric wards, 9 and 4 respectively had nurses trained to perform Ultra using the battery-operated GeneXpert Edge. Laboratory technicians performed the testing in the other sites. We compared proportion of samples tested, invalid or error results, TB detection yield, Turnaround Time (TAT) between sample reception and result at PHC, and between sample collection and result delivery to clinicians in paediatric wards between nurses and laboratory technicians. External Quality Assessment (EQA) and site support supervision assessed performances. Self-administered questionnaire and semi-structured individual interviews assessed nurses’ perceptions.

**Results:** Ultra was done in 253/254 (99.6%) and 258/258 (100%) samples for PHC and hospital nurses vs 895/897 (99.8%) and 874/874 (100%) for laboratory technicians, respectively. At PHC, the TAT was below 1h30 for 158/252 (62.7%) samples tested by nurses vs 677/893 (75.8%) by laboratory technicians, p<0.001. Ultra results were available to clinicians within 3h in 201/258 (77.9%) samples for nurses vs 464/874 (53.1%) for laboratory technicians in hospitals, p<0.001. EQA results <87.5% was more common for PHC nurses than PHC laboratory technicians or hospital nurses. Technical difficulties, lack of practice and workload were the main challenges, and training and supervision the main facilitators reported by nurses.

**Conclusion:** Task shifting of Ultra testing from laboratory technicians to nurses under close supervision could support decentralisation of TB diagnosis and shorten time to treatment decision for very sick patients.

**Key Messages:** *What is already known on this topic:* Centralisation of childhood tuberculosis cares in many high burden and low middle income countries (LMICs) contribute to the important gap of childhood tuberculosis diagnosis. In 2022, WHO recommended the decentralisation of childhood tuberculosis diagnosis to increase access. The rapid tuberculosis molecular XpertMTB/RIF Ultra test recommended by WHO can be deployed at primary health care level but there is still limited data on its use at this level of care and no data for settings without a laboratory. Another diagnostic challenge is the diagnostic delay, which can be fatal in very sick children. There is no data on in-ward Xpert Ultra testing to shorten diagnostic delays in very sick children.

*What this study adds:* As part of the TB-Speed operational research in 6 countries, we trained nurses from primary health centres without laboratory to perform Xpert Ultra tests on nasopharyngeal aspirate sample, using the GeneXpert Edge equipment. With close supervision and sufficient training, task-shifting of NPA Xpert Ultra testing from laboratory technicians to nurses was feasible and well accepted. Similarly, in order to reduce diagnostic delays in very sick children, we trained hospital nurses to perform in-ward Xpert Ultra tests on nasopharyngeal aspirate in 6 countries. Nurses were able to achieve similar performance as laboratory technicians and the time to treatment decision was shorter when Xpert Ultra testing was done by nurses as compared to laboratory technician. To our knowledge this is the first study assessing Xpert Ultra testing by nurses in high tuberculosis burden and LMICs.

*How this study might affect research, practice or policy:* It is our hope that results from this study will foster the development of content and context specific approach of task-shifting to increase access and quality of healthcare services for TB diagnosis within the same labour force especially in LMICs.

## INTRODUCTION

In 2023, of 10.8 million people estimated to have developed tuberculosis (TB) according to the World Health Organization (WHO), 12% (1.25 million) were children. Only 49% of children with TB were notified to WHO by National TB programs (NTP) mainly due to under diagnosis [1]. Challenges in diagnosing childhood TB include the paucibacilliary nature of pulmonary TB in children resulting in low sensitivity of existing microbiological diagnostic tests and the difficulty of obtaining respiratory samples in young children [2,3]. In addition, the centralisation of childhood TB diagnostic services in several high TB incidence and low and middle income countries (LMIC) reduces access to TB care, and thereby contributes to the gap of childhood TB diagnosis [4].

In 2022, to improve diagnosis and access to treatment for childhood TB, WHO recommended 1) decentralising childhood diagnosis services to low-level peripheral health facilities, and 2) improving the quality of the diagnosis approach and specifically the microbiological testing component, by using the rapid molecular Xpert MTB/RIF Ultra Assay (Ultra; Cepheid, Sunnyvale, CA, USA) on stool samples and nasopharyngeal aspirates (NPA) [5]. The automated GeneXpert platform reduces sample manipulation time and biosafety risks and thereby increases the possibility of using the Xpert assay at low levels of healthcare [6]. The single module GeneXpert Edge (G1), a cheaper, heat and dust resistant, more user-friendly and battery-operated platform could be introduced at primary health centre (PHC) levels where there is a lower number of tests to perform daily [7]. In PHCs without laboratory and thus laboratory technicians, nurses could be trained on how to operate the G1 platform and to perform the Ultra assay. Another use for G1 and reason justifying training of nurses to perform Ultra would be to make the Xpert testing available in paediatric wards, to shorten time to diagnosis for highly vulnerable children outside laboratory opening hours.

Apart for simple point-of-care testing such as urine-lipoarabinomannan (LAM) test, there is no literature on task-shifting of microbiological TB diagnostic tests from laboratory technicians to nurses [8,9]. However, previous experiences in other fields of TB – related health care have shown that task-shifting/sharing from workers in specialised or higher cadres to health care workers (HCW) from a different background could address human resource challenges, address unmet health needs, improve health care quality in terms of service delivery and accessibility after short trainings with support supervisions in health services [10 - 13)].

To address the unmet need to improve access to childhood TB microbiological diagnosis, the TB-Speed Decentralisation study implemented Ultra testing on NPA by either nurses or laboratory technicians in children with presumptive TB at PHC level [14]. Similarly, the TB-Speed Pneumonia study that evaluated systematic TB detection in children admitted for severe pneumonia implemented in-wards Ultra testing on NPA by nurses to reduce time to treatment decision [2].

In this study, we compared the feasibility of Ultra testing on NPA in terms of uptake, performance and turnaround times between nurses and laboratory technicians, and we assessed the nurses’ perception and experience of Ultra testing on NPA within the TB-Speed Decentralisation and Pneumonia studies.

## MATERIALS AND METHODS

### Study design, setting and population

We conducted secondary analyses of multi-country, mixed-methods, research data from the TB-Speed Decentralisation and TB-Speed Pneumonia studies. The TB-Speed Decentralization was an operational research study implemented in Cameroon, Cambodia, Uganda, Mozambique, Côte d’Ivoire and Sierra Leone to assess the impact on paediatric TB case detection of the decentralisation of a comprehensive paediatric TB diagnostic package at district hospitals and PHCs in children <15 years with presumptive TB. Ultra testing of NPA was performed by nurses and laboratory technicians in 9 and 14 PHCs respectively (14). The TB-Speed Pneumonia study was a pragmatic stepped-wedge, cluster-randomized trial evaluating the effect of adding systematic TB detection using Ultra on NPA and stools to the WHO standard of care on mortality of children aged 2-59 months admitted for severe pneumonia in 15 tertiary hospitals in Cameroon, Côte d’Ivoire, Cambodia, Uganda, Mozambique and Zambia. Eleven hospitals had Ultra testing performed on NPA by laboratory technicians when rapid testing was possible in a laboratory, and four hospitals by in-ward nurses (2), (Supplementary material, Table S1). In both studies, stool testing by ultra was performed in a laboratory due to specimen processing requirements prior testing. G1 was introduced in PHCs and paediatric wards.

The site selection was jointly agreed upon based on baseline capacity assessment information by the NTPs and the TB-Speed investigators.

The analyses were conducted among 3 different populations: children <15 years old with presumptive TB enrolled at PHC level in the TB-Speed Decentralisation study; children 2 to 59 months old admitted for severe pneumonia enrolled in the intervention arm of the TB-Speed Pneumonia; and all nurses performing Xpert Ultra on NPA in both studies.

### Training and quality assurance of Ultra testing

Nurses and laboratory technicians were trained to perform Ultra on NPA within a 24-hour and a 3-hour turnaround time (TAT) to Ultra results delivery to clinician in the TB-Speed Decentralisation and Pneumonia study, respectively. A 3-5 days theoretical and practical training by each country’s project national laboratory coordinators included Ultra testing of at least 1mL NPA sample according to the manufactures’ instruction using the G1 platform, sample documentation in laboratory registers, issuing of results, and basic equipment maintenance and troubleshooting.

For external quality assurance (EQA) of Ultra testing, dried culture spots (DCS) with inactivated biomimetic controls and negative control test samples manufactured by the accredited SmartSpot Xpert MTB/RIF Ultra EQA Program (SmartSpot Quality, Johannesburg, South Africa) were dispatched yearly to the respective countries with 12 and 8 DCSs per site (4 DCSs per panel) to be analysed systematically by HCWs bi-annually for TB-Speed Decentralization and quarterly for TB-Speed Pneumonia.

In the TB-Speed Decentralisation sites only, site support supervision visits were planned to supervise the decentralisation of the diagnostic package once every month for the first quarter and then quarterly. During each visit, laboratory specialist from the national referral hospitals assessed PHCs’ laboratory technicians’ and nurses’ performance on Ultra testing (through a specific scoring system) and provided technical guidance. In the TB Speed Pneumonia study, Ultra results per site were reviewed monthly from the GeneXpert equipment platforms by the national laboratory coordinator where, error rates (>5%), invalid and no results rates (>2%) called for actions such as re-training or technical intervention.

### Study Endpoints

We assessed the feasibility of Ultra testing according to 4 specific outcomes. Firstly, the uptake and yield of Ultra testing on NPA samples was assessed based on the following indicators: i) Proportion of Xpert Ultra tests performed on successfully collected NPA samples; ii) Proportion of invalid or error Ultra results; iii) MTB detection yield among Ultra valid tests. Secondly, TAT of Ultra testing defined by the measure of time from sample reception to test result (when the test is complete) in the TB-Speed Decentralisation and time from sample collection to test result delivery to clinician in the TB-Speed Pneumonia. Thirdly, the performance of Ultra testing was assessed by the EQA results in both studies and the performance scores from site support supervision reports in the TB-Speed decentralization study. We classified EQA panel result of 87.5% or above as acceptable. The performance indicators for Ultra-testing included: i) Availability of laboratory procedure for Ultra testing; ii) Testing of NPA samples at PHC as planned; iii) Delivery of Ultra results to clinical team within 24 hours; iv) Proper reporting of results in the sample register; v) Proper entry of sample identification number into the GeneXpert software; and vi) Punctual testing of TB samples (on G1). Each indicator was scored using the Likert scale (1-Never, 2-Rarely, 3-Sometimes, 4-Often, 5-Always). The indicators were assessed through real-time observation. In cases where no sample was available for testing, the NPA Ultra testing procedure was simulated, and any indicators that could not be observed were marked as ‘NA’ (Non-Applicable). Lastly, the experience and perceptions of nurses towards performing Ultra test on NPA samples, using the G1 platform, was assessed using self-administered questionnaires in the TB-Speed pneumonia study, and by semi-structured interviews with nurses in both studies. Barriers, facilitators and possibilities for scaling-up and sustaining Ultra testing on NPA performed by nurses were specifically assessed. Nine nurses (3 males and 6 females), median age 32 [31–36] years from 7 sites in Cambodia, Cameroon and Côte d’Ivoire in the Decentralisation study and 16 nurses (3 males and 13 females), median age 39 [36–44] years, from 4 sites in Côte d’Ivoire, Mozambique and Zambia in the Pneumonia study, were interviewed.

### Data collection

To assess the uptake, yield indicators and TAT, data were retrieved from the clinical database of both studies using REDCap and imported into an excel database. EQA panel sample testing results exported by sites from the GeneXpert platform were uploaded to the SmartSpot platform as .CSV files by country’s laboratory coordinator and frame scores were sent back to laboratory coordinators by the SmartSpot team after analysis. All sites’ results were recorded in an excel database (Supplementary material table S2). Performance indicators were collected in paper-based site support supervision forms completed by supervisors on site. Score results were entered into a REDCap database by research assistants.

Self-administered questionnaires were filled anonymously by nurses on tablets directly into a dedicated REDCap database after each sample testing by Ultra in TB-Speed pneumonia study. Semi-structured interviews, following a pre-defined interview guide, were conducted with nurses either face-to-face or on phone (during the COVID-19 pandemic) by trained social science research assistants (SSRAs) in each country, from June to August 2021, for TB Speed Decentralization and from November 2020 to March 2021, for TB Speed Pneumonia. The interviews were recorded with the nurse’s permission, transcribed and translated to English when required and uploaded on a secure server from where it was retrieved for the purpose of this study.

### Data analysis

Quantitative data were presented using percentages, median and interquartile range (IQR) values by study and by site for EQA and performance scores. Chi-square tests and non-parametric tests were used to compare proportion and median of uptake proportions, TATs and performance score results between nurses and laboratory technicians by study using R studio software (version 4.3.2, R Foundation for Statistical Computing).

For qualitative data, codebooks were developed by the implementation research team, based on deductive and data led approach. Interviews were coded by country SSRAs and analysed by country in Nvivo Release qualitative data management software, QSR International Private Ltd. Version 1.5, 2021. Multi-country intercoder reliability was maximized by SSRAs coding the same three transcripts and reviewing together the coded data. Thematic summaries were prepared by SSRAs, with support from the study investigators; themes were defined based on the Sekhon theoretical framework of acceptability where we categorized themes under relevant domains of the framework as follows: Knowledge; Burden (workload, technical issues, long and difficult sample process, lack of samples); and Self-efficacy (training, supervision visits, support from research team, specific delegation of task) [15].

### Ethical statement

The study was approved by National Ethics Committees of all the participating countries, the WHO Ethical Review Board, and the Institutional Review Board of Inserm (Institute National de la Santé et de la Recherche Médicale, Paris, France). Individual consent was obtained from parent/guardian of each child before enrolment in the studies and an assent was obtained if the child was old enough. Written informed consent was taken prior administration of questionnaires and interviews from all the nurses who participated.

## RESULTS

In terms of uptake, at PHC level, 253/254 (99.6%) and 895/897 (99.8%) samples were tested by nurses and laboratory technicians respectively, and at referral hospitals, all samples were tested. There was no significant difference of detection yield between nurses and laboratory technicians. Although the proportion of invalid and error results was overall very low (< 5%) it was significantly higher when testing was done by nurses vs laboratory technicians at both PHC and referral hospitals (Table 1).

**Table 1:** Uptake of Xpert Ultra testing on nasopharyngeal aspirate by nurses and laboratory technicians.

|  | Primary Health Centres |  |  | Reference hospitals |  |  |
| --- | --- | --- | --- | --- | --- | --- |
|  | Nurses | Laboratory technicians | P value | Nurses | Laboratory technicians | P value |
| Samples received, N | 254 | 897 |  | 258 | 874 |  |
| Samples tested, n (%) | 253 (99.6)* | 895 (99.8)* | 0.637 | 258 (100) | 874 (100) | NA |
| MTB detected, n (%) | 2 (0.8) | 3 (0.3) | 0.348 | 1 (0.4) | 20 (2.3) | 0.047 |
| MTB not detected, n (%) | 242 (95.7) | 877 (98.0) | 0.037 | 249 (96.5) | 847 (96.9) | 0.748 |
| Invalid or Error, n (%) | 9 (3.6) | 11 (1.2) | 0.012 | 8 (3.1) | 7 (0.8) | 0.005 |

MTB: *Mycobacterium tuberculosis*; NA: non applicable
*\*1 missing sample by nurses and 2 missing samples by laboratory technicians due to technical issues and sample not received by the laboratory.*

All PHCs participated in both EQA panels but one due to a missing EQA panel. There were 2/9 (33.3%) PHCs with nurses that had results > 87.5% in all EQA panels vs 9/14 (64.3%) for PHC with laboratory technicians, p=0.487. At referral hospital, 1/4 sites with Ultra testing done by nurse and 3/11 sites with testing done by laboratory technician did not participate in any EQA panel because panels had expired when the sites were randomized to the intervention. Since the TB-Speed pneumonia study had a stepped wedge design, the number of EQA panels per site depended on the time of randomization to the intervention of each site. There were 2/3 (66.7%) sites that had all EQA panel results > 87.5% vs 6/8 (75.0%) in sites with nurses or laboratory technicians (P=0.640), respectively (Figure 1).

**Figure 1.**
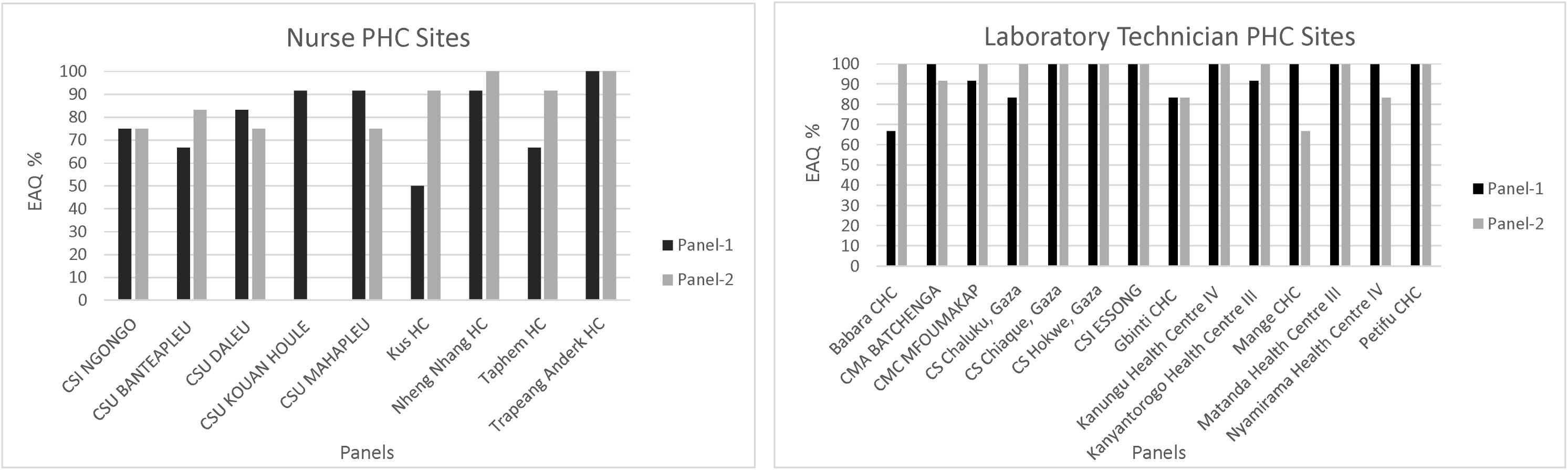
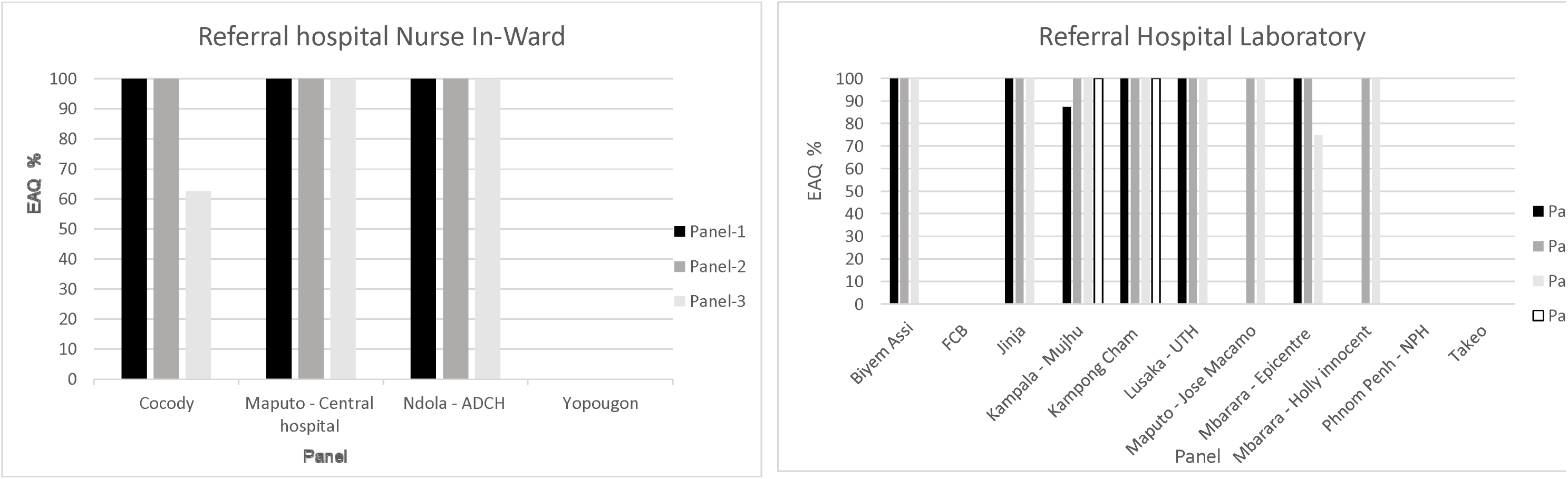
External Quality Assurance Panel results Figure 1a: For nurses at Primary Health Centre sites Figure 1b: For laboratory technicians at Primary Health Centre sites Figure 1c: For nurses at referral hospitals Figure 1d: For laboratory technicians at referral hospitals

The median TAT of Ultra testing from sample reception to Ultra results at PHC levels, was 1.13 hour (h) [IQR 1.08; 1.79] for nurses and 1.10h [1.07; 1.45] for laboratory technicians, p < 0.001. The TAT was below 1h30 for 158/252 (62.7%) samples tested by nurses vs 677/893 (75.8%) for samples tested by laboratory technicians, p<0.001. At referral hospitals, the TAT from sample collection to results delivery was 1.95h [1.5; 2.87] for nurses and 2.87h [1.97; 4.81] for laboratory technicians, p <0.001. The TAT was within 3h for 201/258 (77.9%) samples for nurses vs 464/874 (53.1%) for laboratory technicians, p<0.001.

The median performance score results ranged between 4 and 5 out of 5 points for all performance indicators at PHC levels for sites with nurses and laboratory technicians (Table 2).

**Table 2:** Performance score results of site support supervision at primary health care levels.

| Site | Number of visits | Indicator 1<br>Median<br>(Q1 – Q3) | Indicator 2<br>Median<br>(Q1 – Q3) | Indicator 3<br>Median<br>(Q1 – Q3) | Indicator 4<br>Median<br>(Q1 – Q3) | Indicator 5<br>Median<br>(Q1 – Q3) | Indicator 6<br>Median<br>(Q1 – Q3) |
| --- | --- | --- | --- | --- | --- | --- | --- |
| **Laboratory technicians** |  |  |  |  |  |  |  |
| Sierra Leone - Babara | 4 | 5 (4.75 - 5) | 5 (5 - 5) | 5 (5 - 5) | 4 (4 - 4.5) | 4 (4 - 4.5) | 5 (5 - 5) |
| Cameroon - Batchenga | 5 | 5 (5 - 5) | 5 (4.5 - 5) | 5 (5 - 5) | 5 (5 - 5) | 5 (5 - 5) | 5 (4.75 - 5) |
| Cameroon - Mfomakap | 3 | 5 (3.5 - 5) | 5 (5 - 5) | 5 (5 - 5) | 5 (5 - 5) | 5 (5 - 5) | 5 (5 - 5) |
| Cameroon - Essong | 5 | 5 (5 - 5) | 5 (5 - 5) | 5 (5 - 5) | 5 (5 - 5) | 5 (5 - 5) | 5 (5 - 5) |
| Mozambique - Chalucwane | 4 | 5 (5 - 5) | 5 (5 - 5) | 4.5 (4 - 5) | 4.5 (4 - 5) | 5 (4.75 - 5) | 4 (4 - 4.25) |
| Mozambique - Chiaqelane | 4 | 5 (5 - 5) | 5 (5 - 5) | 5 (4.75 - 5) | 4.5 (3.25 - 5) | 5 (4.25 - 5) | 4.25 (4 - 5) |
| Sierra Leone - Gibinti | 5 | 5 (5 - 5) | 5 (5 - 5) | 5 (5 - 5) | 5 (5 - 5) | 4.5 (4 - 5) | 5 (4.75 - 5) |
| Mozambique - Hokwe | 4 | 5 (5 - 5) | 5 (5 - 5) | 5 (5 - 5) | 5 (5 - 5) | 4.5 (4 - 5) | 5 (5 - 5) |
| Uganda - Kanungu | 5 | 5 (5 - 5) | 5 (5 - 5) | 4 (4 - 4) | 5 (5 - 5) | 5 (5 - 5) | 3 (3 - 4) |
| Uganda - anyantorogo | 5 | 5 (5 - 5) | 5 (5 - 5) | 5 (5 - 5) | 5 (5 - 5) | 5 (5 - 5) | 5 (4.75 - 5) |
| Sierra Leone - Mange | 4 | 5 (4.75 - 5) | 5 (4.25 - 5) | 5 (5 - 5) | 4.5 (3.75 - 5) | 4 (4 - 4.5) | 5 (4.5 - 5) |
| Uganda - Matanda | 5 | 5 (5 - 5) | 5 (5 - 5) | 5 (5 - 5) | 5 (5 - 5) | 5 (5 - 5) | 5 (5 - 5) |
| Uganda - Nyamirama | 5 | 5 (5 - 5) | 5 (5 - 5) | 5 (5 - 5) | 5 (5 - 5) | 5 (5 - 5) | 5 (5 - 5) |
| Sierra Leone - Petifu | 5 | 5 (5 - 5) | 5 (4 - 5) | 5 (2 - 5) | 5 (4 - 5) | 5 (4 - 5) | 5 (5 - 5) |
| **Nurses** |  |  |  |  |  |  |  |
| Site | Number of visits | Indicator 1<br>Median<br>(Q1 – Q3) | Indicator 2<br>Median<br>(Q1 – Q3) | Indicator 3<br>Median<br>(Q1 – Q3) | Indicator 4<br>Median<br>(Q1 – Q3) | Indicator 5<br>Median<br>(Q1 – Q3) | Indicator 6<br>Median<br>(Q1 – Q3) |
| Cambodia - Kus | 4 | 5 (5 - 5) | 4 (4 - 4) | 5 (5 - 5) | 4 (3.5 - 4.5) | 4.5 (4.25 - 4.75) | 5 (5 - 5) |
| Cameroon - Ngongo | 5 | 5 (5 - 5) | 5 (5 - 5) | 5 (5 - 5) | 5 (5 - 5) | 5 (5 - 5) | 5 (5 - 5) |
| Cambodia - Ta Phem | 4 | 5 (5 - 5) | 5 (5 - 5) | 5 (5 - 5) | 4 (3 - 4.5) | 4 (4 - 4.5) | 5 (5 - 5) |
| Cambodia - Nheng Nhang | 4 | 5 (5 - 5) | 5 (5 - 5) | 5 (5 - 5) | 5 (5 - 5) | 5 (5 - 5) | 5 (5 - 5) |
| Cambodia - Tropang Andert | 4 | 5 (5 - 5) | 5 (5 - 5) | 5 (5 - 5) | 4.5 (4.25 - 4.75) | 5 (5 - 5) | 5 (5 - 5) |

**IQR: Inter quartile range**
**Likert scale: 1-Never, 2-Rarely, 3-Sometimes, 4-Often, 5-Always, and NA**
**Indicator 1** = Availability of Laboratory SOPs for TB testing
**Indicator 2** = Testing of NPA samples for TB on site
**Indicator 3** = NPA results sent to clinical team within 24 hours
**Indicator 4** = Proper reporting of results in the specimen register
**Indicator 5** = Proper entry of sample IDs into the GeneXpert software
**Indicator 6** = Timely testing of TB samples (on GeneXpert)

**Table 3:** Self-administered questionnaires on Xpert Ultra testing from nasopharyngeal aspirate reported by nurses at referral hospital level.

|  | **NPA samples tested (N=156)** |
| --- | --- |
| **How would you qualify the Ultra test you performed for this child?** |  |
| Easy | 72 (46.2) |
| Neither easy nor difficult | 73 (46.8) |
| Difficult | 6 (3.8) |
| Impossible | 4 (2.6) |
| Missing | 1 (0.6) |

| Did you face any technical difficulties during this Ultra testing? |  |
| --- | --- |
| No | 137 (87.8) |
| Yes | 18 (11.5) |
| Missing | 1 (0.6) |
| **Technical difficulties reported by study nurse running Ultra on G1** | **Nurses (N=18)** |
| Spilling of sample occurred | 0 (0) |
| Issue while pipetting the sample or the reagent | 3 (16.7) |
| Difficulties in manipulating the G1 edge | 0 (0) |
| Technical error occurred when running the test | 14 (77.8) |
| Ran out of battery | 1 (5.6) |
| Unidentified reason for the technical difficulty | 0 (0) |
| Other | 2 (11.1) |

NPA: Nasopharyngeal Aspirate

Difficulties were mostly related to the occurrence of technical errors when running the tests. During interviews, nurses reported that, during the first few weeks, performing Ultra testing was not so easy, but that they had felt more confident with time and practice*: “First, the process is difficult, long process for it. For other work process, if we do more and more, we get faster, but for this process, it is related to a lot technical, like computer and typing. It takes time it is difficult” (Cambodia-PHC Nurse).* Among the challenges, nurses reported experiencing technical issues with scanning barcode of the cartridge: *“Sometimes, scanning barcode doesn’t match with cartridge. So, the cartridge will be broken, and we need to change new one. When scanning doesn’t match, we disconnect the cable from computer and then connect and scan again. Sometime, we put it in machine but computer also errors. It ran for a while then it can run backward. I had met this challenge. Then, I tried to put it again and call to Mr. XXXX and he suggested to put it again” (Cambodia-PHC Nurse);* inserting the cartridge in the GeneXpert module: *“With the machine, what we often have difficulty with is the last stage where the machine has to open, the part where you put the cassette in, and it often refuses, so you have to start again several times. (…) So that’s the difficulty. Otherwise, when it works, there are no problems” (Cote d’Ivoire PHC Nurse);* and using the computer for documentation and navigating the platform: *“My dear, we don’t know how to handle computers, that’s our problem (laughs). We don’t know how to handle computers. I myself had to…it’s because I had to force it…force it a little bit, that’s my little worry, technology is not my forte. But as I wanted. I was curious, so I forced it” (Cote d’Ivoire, ward nurse*).) Another challenge more specific to PHC nurses was the lack of practice, due to few samples to test each week, “*If I did not do it long time, I will forget it again. … I get confused. If I do it more often I will remember it.” (Cambodia-PHC nurse).* On the other hand, some nurses reported the issue of increase in workload, especially when they have to re-run a test due to error result: “*….it’s an extra activity yeah, …. it’s the lab to run except the RDT, now in this case it me to wait for probably 87 minutes for it to run, if it gives me error then I need to retrieve the bio-banked sample I start preparing, I has given me a work overload” (Zambia, Ward Nurse)*. Nurses highlighted several experiences of facilitators that included the training and training materials that they could consult after training and the support during supervision visits: *“When I take the NPA sample, I mark everything because you have to mark the time of sampling. I go to my laboratory because it’s a small lab there, I go to the lab, the procedure for doing the technique is there, I look at the procedure, I technique the NPA and then I put it into the Xpert and I wait 87 minutes and then the result comes out.”* (*Cote d’Ivoire PHC Nurse).* Nurse explained that the technical challenges could be overcome by contacting laboratory technicians: “*Sometimes when we have such errors we usually call for a meeting with the lab so that we help and understand why we are having such errors, so they usually attend.” (Zambia, ward nurse)*. They appreciated the battery operated equipment: *“(…) the machine itself runs on an autonomous battery, and we have up to two batteries. So even if there’s a power cut, we can work”.* (*Cote d’Ivoire PHC Nurse).* Additional results can be found in Supplementary material Table S3.

## DISCUSSION

In these two studies, task-shifting of Ultra testing on NPA samples with G1 from laboratory technicians to nurses was feasible and well appreciated irrespective of the level of facility with the support of training, supervision and quality assurance. These findings support the decentralization of childhood TB diagnosis at PHC level to increase access to care [16]. Using Ultra testing by nurses in-ward significantly reduced the time to treatment decision as compared to sites where testing was done in laboratory by laboratory technician.

The uptake of Ultra testing was very high in all facilities with either nurse or laboratory technician with only few invalid or error results. However, the proportion of invalid or error results was higher when testing was done by nurses, which is consistent with the lower EQA results in sites where the testing was done by nurses, especially at PHC level. This highlights the importance of training and supervision, as raised by the nurses during the interviews. The PHC nurses mentioned their challenge with the lack of experience due to the few samples to be tested per week, which can explain the trend of lower performance by PHC nurses than by referral hospital nurses. The number of tests per week might be a criterion to consider before decentralising Ultra testing at PHC level to secure good performance.

Among challenges, nurses mostly reported technical challenges in relation to laboratory practices such as pipetting or to data entry. Increased workload, fear of getting infected from positive samples, and lack of reliable power supply were additional challenges reported by nurses, which are similar to previous xperiences regarding implementation of on-site molecular TB testing in Uganda and Mongolia [17, 18]

Training and close supervision were highlighted as key facilitators by nurses. This is consistent with previous experience on task shifting of TB–HIV co-infection management in Uganda [19].

In as-much-as task-shifting was possible in our study, we need to be weary of the workload of the nurses and to recommend that a content and context-specific approach should be considered for task shifting where applicable to support nurses to effectively adapt to performing new tasks.

Within TB-Speed, Ultra testing was a new experience for nurses and was met with enthusiasm about the positive impact it would have on the quality of childhood TB diagnosis services. The experience of learning something technical and independently executing it after a few days of training and periodic support gave the nurses a sense of confidence, increased their knowledge and autonomy in their capacity to provide reliable and quality health care services in shorter time to the community. This tends to increasing job satisfaction of the HCWs, [18,20]. Nurses at PHC levels perceived that being able to provide such a high level of service which is normally not at their level to provide, will boost their career, increase confidence of the community in them and the visibility of their health facilities and roll-out patient referral, as mentioned in the interviews, thus, increasing population access and coverage, [18, 20–22].

The main study limitation was the potential site selection bias. Sites with Ultra testing done by nurses or laboratory technicians were selected for convenience based on implementation requirements following site assessment. Therefore, sites implementing Ultra by nurses or laboratory technicians were not randomly selected. Therefore, we did not control for the heterogeneity between sites and it is possible that other factors related to the sites independent of the personnel performing the testing could affect the study endpoints. Although it was an operational research done with existing facility staff, the study conditions in terms of support may not reflect what would be available in real life.

In conclusion, the study findings suggest that task-shifting from laboratory technicians to nurses for on-site TB molecular diagnostic testing could contribute to improve access to microbiological TB diagnosis at PHCs and could reduce time to treatment decision. Regardless of the technological advancements and development of molecular testing platforms geared towards increasing accessibility to tests and overcoming infrastructural and environmental limitations, close support supervision and support after training is key in identifying and addressing underlying challenges. This can aid to re-design or develop context and content-specific training programs followed by periodic refresher trainings to build the capacity of the HCW in adopting and performing new tasks [18, 20–22]. Human resource shortage and high mobility are a huge challenge in LMICs and task-shifting could contribute to fill-in this gap.

## Data Availability

According to the French and European legal restrictions law no. 78-17 of January 6, 1978 called Loi informatique et liberte and European law GDPR General Data Protection Regulation that govern data sharing, data cannot be shared to an undeclared third party if study participants have not consented. Data could be made available by the sponsor (Inserm) to any researcher interested under a data transfer agreement to the study coordinating investigator or to the sponsor

## ACKNOWLEGMENTS

We thank the Ministries of Health and National Tuberculosis Programs of participating countries for their support. We thank all the children and their families who participated to the study and the healthcare workers of the participating health facilities. We thank the members of the TB-Speed Scientific Advisory Board who gave technical advice on the design of the study: Steve Graham (The University of Melbourne, Melbourne, Australia), Anneke Hesseling (Stellenbosch University, Cape Town, South Africa), Luis Cuevas (Liverpool School of Tropical Medicine, Liverpool, UK), Christophe Delacourt, Sabine Verkuijl (WHO, Geneva, Switzerland), Philippa Musoke (Makerere University, Kampala, Uganda), Elizabeth Maleche-Obimbo (University of Nairobi, Kenya) Mao Tan Eang (CENAT, Cambodia). Findings from this study were presented during the 54^th^ Union World Conference on Lung Health, that was held on November 15–18, 2023 in Paris.

## AUTHORS’ CONTRIBUTION

OM, MB, EW conceived and designed the TB-Speed Decentralisation and Pneumonia studies. JOG coordinated the qualitative component of both studies. OM, MB, and EW led the study at international level. JVT, RM, GB, JMA, LB, CC, CK and EW coordinated the implementation of these studies in the different countries. RK, MN, KK, EM, SY, NEM, TO and JM coordinated laboratory activities at country level in the different countries. BB and BJ coordinated on site qualitative data collection. ML and NEM coordinated laboratory activities at the international level. NEM as part of here PHD project collected data, performed analyses and wrote the first draft of the manuscript with support from JOG, MB, BB and BJ. All authors reviewed and approved the final version of the manuscript.

## REFERENCE

1. Global Tuberculosis Report 2024 [Online]. 2025. Available from: https://www.who.int/teams/global-programme-on-tuberculosis-and-lung-health/tb-reports/global-tuberculosis-report-2024 (accessed 18 March 2025)

2. Marcy O, Wobudeya E, Font H, et al. Effect of systematic tuberculosis detection on mortality in young children with severe pneumonia in countries with high incidence of tuberculosis: a stepped-wedge cluster-randomised trial. Lancet Infect Dis. 2023;23:341–51.

3. Brown S, Leavy JE, Jancey J. Implementation of GeneXpert for TB Testing in Low- and Middle-Income Countries: A Systematic Review. Glob Health Sci Pract. 2021 9:698–710.

4. Wobudeya E, Bonnet M, Walters EG, et al. Diagnostic Advances in Childhood Tuberculosis—Improving Specimen Collection and Yield of Microbiological Diagnosis for Intrathoracic Tuberculosis. Pathogens. 2022;11:389.

5. WHO operational handbook on tuberculosis: module 5: management of tuberculosis in children and adolescents [Internet]. [Online 2025]. Available from: https://www.who.int/publications/i/item/9789240046832 (accessed 20 March 2025)

6. Kay AW, Ness T, Verkuijl SE, et al. Xpert MTB/RIF Ultra assay for tuberculosis disease and rifampicin resistance in children. Cochrane Database Syst Rev. 2022:CD013359.

7. Reza TF, Nalugwa T, Farr K, et al. Study protocol: a cluster randomized trial to evaluate the effectiveness and implementation of onsite GeneXpert testing at community health centers in Uganda (XPEL-TB). Implement Sci IS. 2020;15:24.

8. Åhsberg J, Tersbøl BP, Puplampu P, et al. Use of the urine Determine LAM test in the context of tuberculosis diagnosis among inpatients with HIV in Ghana: a mixed methods study. Front Public Health. 2023;11:1271763.

9. Peter JG, Zijenah LS, Chanda D, et al. Effect on mortality of point-of-care, urine-based lipoarabinomannan testing to guide tuberculosis treatment initiation in HIV-positive hospital inpatients: a pragmatic, parallel-group, multicountry, open-label, randomised controlled trial. Lancet Lond Engl. 2016;387:1187–97.

10. Naikoba S, Senjovu KD, Mugabe P, et al. Improved HIV and TB Knowledge and Competence Among Mid-level Providers in a Cluster-Randomized Trial of One-on-One Mentorship for Task Shifting. JAIDS J Acquir Immune Defic Syndr. 2017;75:e120–7.

11. Peresu E, Heunis JC, Kigozi NG, et al. Task-shifting directly observed treatment and multidrug-resistant tuberculosis injection administration to lay health workers: stakeholder perceptions in rural Eswatini. Hum Resour Health. 2020;18:97.

12. Mafigiri DK, McGrath, Janet W., et al. Task shifting for tuberculosis control: A qualitative study of community-based directly observed therapy in urban Uganda. Glob Public Health. 2012;7:270–84.

13. Farley JE, Ndjeka N, Kelly AM, et al. Evaluation of a nurse practitioner-physician task-sharing model for multidrug-resistant tuberculosis in South Africa. PLOS ONE. 2017;12:e0182780.

14. Wobudeya E, Nanfuka M, Ton Nu Nguyet MH, et al. Effect of decentralising childhood tuberculosis diagnosis to primary health centre versus district hospital levels on disease detection in children from six high tuberculosis incidence countries: an operational research, pre-post intervention study. eClinicalMedicine. 2024;70:102527.

15. Sekhon M, Cartwright M, Francis JJ. Development of a theory-informed questionnaire to assess the acceptability of healthcare interventions. BMC Health Serv Res. 2022;22:279.

16. Joshi B, De Lima YV, Massom DM, et al. Acceptability of decentralizing childhood tuberculosis diagnosis in low-income countries with high tuberculosis incidence: Experiences and perceptions from health care workers in Sub-Saharan Africa and South-East Asia. Robinson J, editor. PLOS Glob Public Health. 2023;3:e0001525.

17. Rendell NL, Bekhbat S, Ganbaatar G, et al. Implementation of the Xpert MTB/RIF assay for tuberculosis in Mongolia: a qualitative exploration of barriers and enablers. PeerJ. 2017 ;5:e3567.

18. Nalugwa T, Handley M, Shete P, et al. Readiness to implement on-site molecular testing for tuberculosis in community health centers in Uganda. Implement Sci Commun. 2022;3:9.

19. Senjovu DK, Naikoba S, Mugabe P, et al. Retention of knowledge and clinical competence among Ugandan mid-level health providers 1 year after intensive clinical mentorship in TB and HIV management. Hum Resour Health. 2021;19:150.

20. Okoroafor SC, Christmals CD. Task Shifting and Task Sharing Implementation in Africa: A Scoping Review on Rationale and Scope. Healthcare. 2023;11:1200.

21. Okoroafor SC, Dela Christmals C. Health Professions Education Strategies for Enhancing Capacity for Task-Shifting and Task-Sharing Implementation in Africa: A Scoping Review. J Contin Educ Health Prof. 2023 8;

22. Okoroafor SC, Christmals CD. Optimizing the roles of health workers to improve access to health services in Africa: an implementation framework for task shifting and sharing for policy and practice. BMC Health Serv Res. 2023;23:843.

